# Prediction of deterioration from COVID-19 in patients in skilled nursing facilities using wearable and contact-free devices: a feasibility study

**DOI:** 10.1101/2022.04.06.22273202

**Authors:** Sabine von Preyss-Friedman, Linda Emmet, Dree Deckert, Dennis A.B. David, Heikki Raisanen, Kevin Longoria, Willem Gielen, Martin G. Frasch

**Affiliations:** Avalon Healthcare, Salt Lake City, Utah, UT, USA; Health Stream Analytics LLC, Seattle, WA, USA; Emfit Ltd, Finland, USA; Biostrap, Duarte, CA, 91010, United States

**Author notes:** **Corresponding author:** Dr. Martin Frasch, Vitalink AI Inc, 7724 35th Ave NE, #15897, Seattle, WA 98115.

## Abstract

**Background and Rationale:** Approximately 35% of all COVID-19 deaths occurred in Skilled Nursing Facilities (SNFs). In a healthy general population, wearables have shown promise in providing early alerts for actionable interventions during the pandemic. We tested this promise in a cohort of SNFs patients diagnosed with COVID-19 and admitted for post-acute care under quarantine. We tested if 1) deployment of wearables and contact-free biosensors is feasible in the setting of SNFs and 2) they can provide early and actionable insights into deterioration.

**Methods:** This prospective clinical trial has been IRB-approved (NCT04548895). We deployed two commercially available devices detecting continuously every 2-3 minutes heart rate (HR), respiratory rate (RR) and uniquely providing the following biometrics: 1) the wrist-worn bracelet by Biostrap yielded continuous oxygen saturation (O2Sat), 2) the under-mattress ballistocardiography sensor by Emfit tracked in-bed activity, tossing, and sleep disturbances. Patients also underwent routine monitoring by staff every 2-4 h. For death outcomes, cases are reported due to the small sample size. For palliative care versus at-home discharges, we report mean±SD at p<0.05.

**Results:** From 12/2020 - 03/2021, we approached 26 PCR-confirmed SarsCoV2-positive patients at two SNFs: 5 declined, 21 were enrolled into monitoring by both sensors (female=13, male=8; age 77.2±9.1). We recorded outcomes as discharged to home (8, 38%), palliative care (9, 43%) or death (4, 19%). The O2Sat threshold of 91% alerted for intervention. Biostrap captured hypoxic events below 91% nine times as often as the routine intermittent pulse oximetry. In the patient deceased, two weeks prior we observed a wide range of O2Sat values (65-95%) captured by the Biostrap device and not noticeable with the routine vital sign spot checks. In this patient, the Emfit sensor yielded a markedly reduced RR (7/min) in contrast to 18/min from two routine spot checks performed in the same period of observation as well as compared to the seven patients discharged home over a total of 86 days of monitoring (RR 19± 4). Among the patients discharged to palliative care, a total of 76 days were monitored, HR did not differ compared to the patients discharged home (68±8 vs 70±7 bpm). However, we observed a statistically significant reduction of RR at 16±4/min as well as the variances in RR (10±6 vs 19±4/min vs16±13) and activity of palliative care patients vs. patients discharged home.

**Conclusion/Discussion:** We demonstrate that wearables and under-mattress sensors can be integrated successfully into the SNF workflows and are well tolerated by the patients. Moreover, specific early changes of oxygen saturation fluctuations and other biometrics herald deterioration from COVID-19 two weeks in advance and evaded detection without the devices. Wearable devices and under-mattress sensors in SNFs hold significant potential for early disease detection.

## Introduction

Across the U.S., the population of 55 million seniors, persons 65 and older, has been the hardest hit by COVID-19 - especially those living in long-term care assisted living and post-acute care (LTC) facilities and nursing homes (**Figure 1**).

**Figure 1.**
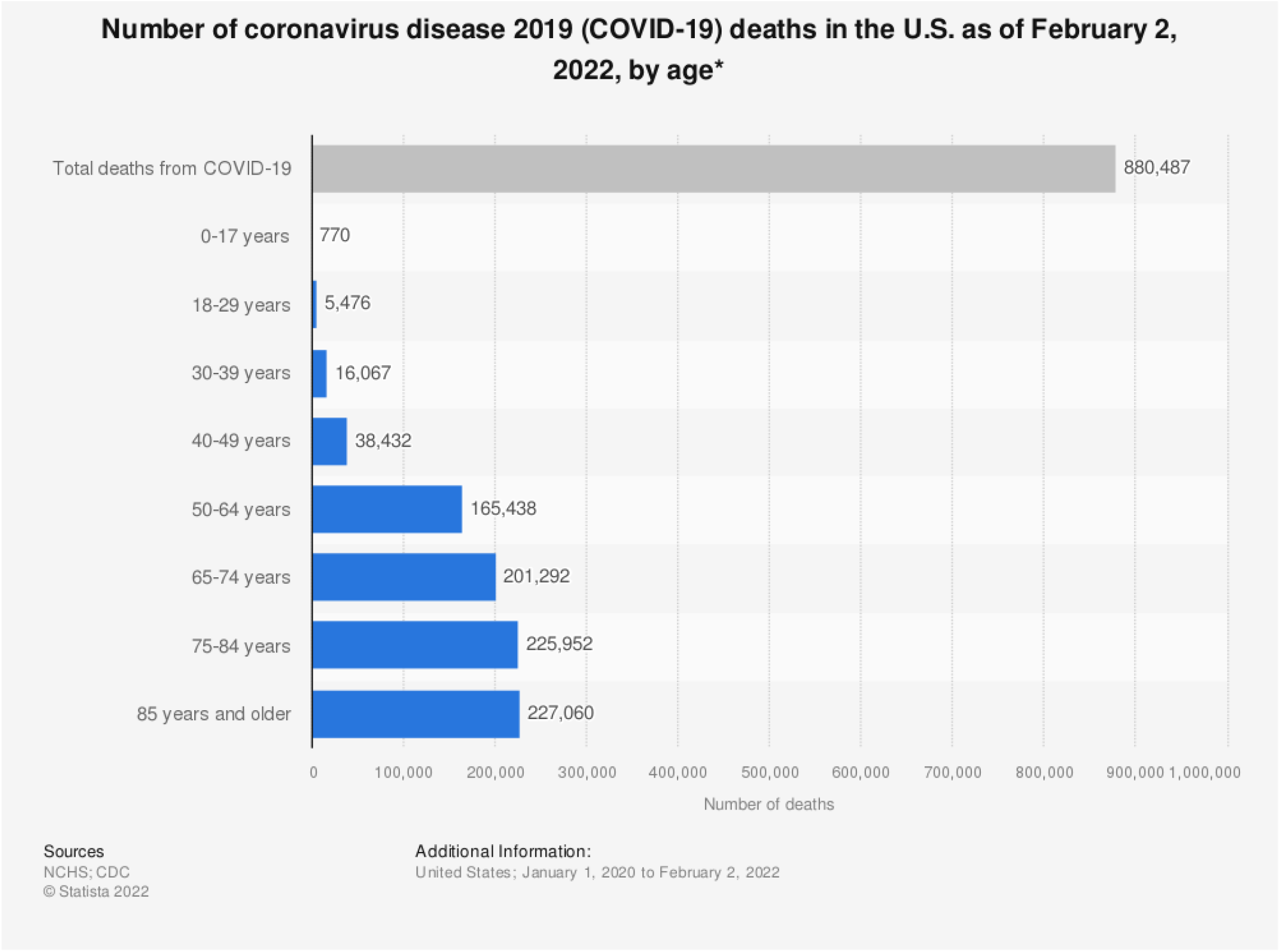
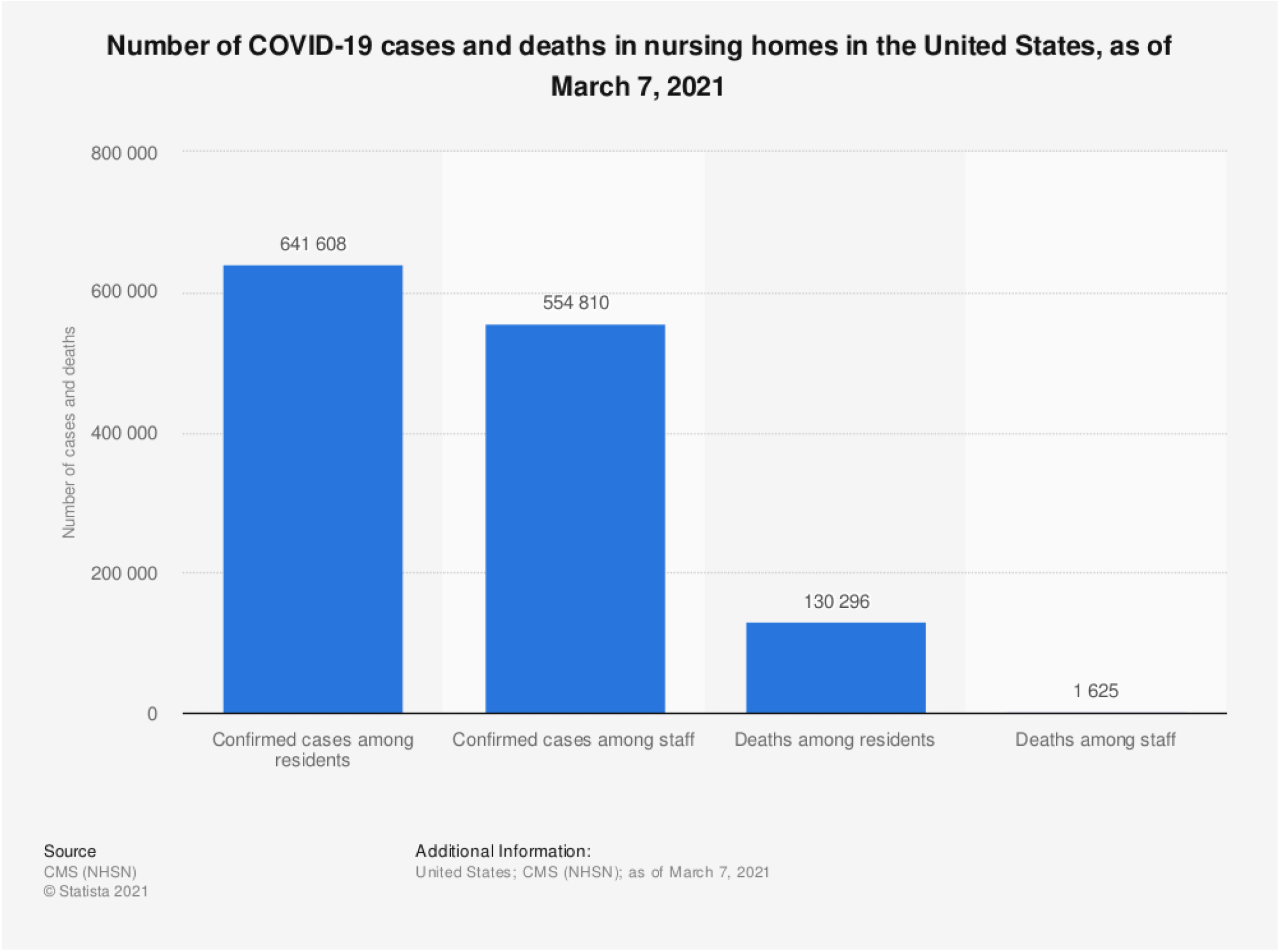
Impact of COVID-19 on the senior population in the U.S. **TOP. 74% of COVID-19 deaths in the United States were in the people 65 and older**. Data as of February 2, 2022, surveyed from January 1, 2020.[1] **BOTTOM**. As of March 7, 2021, there had been a total number of 641,608 confirmed COVID-19 cases and 130,296 deaths among nursing home residents in the United States. The number of COVID-19 cases among nursing home staff in the United States reached 130,296 cases, as of March 7, 2021.

Patients with a mild illness caused by COVID-19 can deteriorate unexpectedly, require intensive care or die from respiratory distress or cardiac arrest. As of February 2, 2021, The New York Times reported the case-fatality rate among nursing home residents to be 10%, five times the average general population case-fatality rate.[2] The virus has infected more than 1,234,000 people at 31,000 LTC facilities and killed over 152,000 LTC residents. In the U.S., 654,304 deaths occurred among people over 65 years of age which is 74% of all deaths as of October 20, 2021 (Figure 1). LTC-associated cases accounted for only 5% of detected infections but 34% of all deaths from COVID-19. In 11 US states, at least half of COVID-19 deaths were linked to nursing homes, because spread there put frontline healthcare workers and their families at risk of contracting the virus.[2] In U.K., in a recent and largest study of COVID-19 in a population of 17,3 million adults [3], it was reported that over 90% of UK deaths were in people over 60. The statistics in the US echoes this finding. As the pandemics are bound to reoccur and the population of 65 years and older continues to rise, projected to reach 22% of U.S. total population by 2050, the urgency the present virus spread has highlighted will also continue to rise.[4]

As of today, there has not been any coordinated efforts to help this underserved population systematically and long-term to prevent these dramatic mortality rates from reoccurring. Despite the recent advances to vaccinate seniors as a priority, booster shots will continue to be required and hundreds of thousands of senior citizens remain at exponentially higher risk of dying from this disease or similar conditions causing respiratory distress. In addition to the immediate death toll, the pandemic has highlighted long-standing inefficiencies in delivering healthcare to this population, wherein serious infections are often missed in their subclinical phases until irreversible, catastrophic organ damage makes effective treatment too late.

Due to inherent infrastructural limitations, the critical vital signs (e.g., oxygen saturation, heart rate, respiratory rate) measurements in LTCs are not obtained frequently enough (every four hours tends to be the maximum frequency feasible during pandemic emergencies). This may lead to missed signs of deterioration.

Patients with a mild illness caused by COVID-19 can deteriorate unexpectedly. Once a mild infection has been diagnosed, most patients are sent home to self-quarantine, but about 15% of all adults infected with COVID-19 deteriorate without sufficient warning and require intensive care or and 4.6% overall die from respiratory distress or cardiac arrest. That risk is over 3-fold of the general population or 10% among the residents of nursing homes/LTACs (as cited above).

In a healthy general population, wearable devices have shown promise in providing early alerts for actionable interventions during the pandemic.[5,6]

Consequently, the first objective of the present study was to test if 1) deployment of wearables and contact-free biosensors is feasible in the setting of SNFs and 2) they can provide early and actionable insights into deterioration from COVID-19.

## Methods

This prospective clinical trial entitled “**Nursing Homes and COVID-19”** has been approved by the Western IRB (**Protocol Number: HSA-001) and** registered on clinicaltrials.gov as NCT04548895.

The presented findings are derived from the study on two COVID-19 units of LTC facilities deploying two commercially available biomonitoring technologies which detected heart rate and computed the respiration as well as uniquely provided the following biometrics:

1) the wrist-worn bracelet biosensors by Biostrap yielded continuous clinical-grade oxygen saturation, and
2) the under-mattress ballistocardiography (BCG) sensors by Emfit tracked in-bed activity, tossing and turning, and sleep disturbances.

### Study Design

#### Phase

This is an ecologic proof-of-concept study.

#### Setting

The study took place in two skilled nursing facilities providing long-term post-acute care (LTCF) in the state of Washington in the United States (henceforth collectively referred to as “LTCF”).

#### Interventions

##### Biometric monitors – wearable and under-the-mattress

LTCF residents in settings with anticipated high COVID-19 incidence, who agreed to participate in the study and completed the informed consent process were each given a bracelet-like wearable and/or an under-the-mattress monitor. The study was conducted in collaboration with Biostrap providing its wristband wearable and Emfit providing the under-mattress sensor. Both companies also provided their respective de-identified cloud services to capture and analyze the data.

Emfit devices were installed once after enrollment under each participant’s mattress and left plugged in and connected to the cloud via WiFi to record automatically without further intervention. The participants wore their wristbands consistently, ideally 24 hours a day, 7 days a week, for the duration of their stay in the COVID-19 unit. The wristbands transferred data continuously and wirelessly to a tablet via Bluetooth (without adding resident-to-staff contact) from the tablet via WiFi to the cloud. Nursing assistants ensured the Biopstrap devices were charged wirelessly on their docking stations once a day for 20 minutes during afternoon meals or patient care periods.

#### Monitor output

The photoplethysmography (PPG)-based Biostrap wristbands recorded heart rate, computed respiratory activity, oxygen saturation, and sleep/activity patterns continuously. The ballistocardiography (BCG)-based under-mattress (i.e., contact-free) Emfit sensor captured approximately every 10 seconds HR, RR and movements of the patients while they were in bed.

For the purposes of this project, Health Stream Analytics downloaded the raw data from each participant’s manufacturer-hosted cloud account for offline analysis. The real-time monitoring implementation provides live data analytics.

The standard of care was not affected by the study.

#### Questionnaires and vital signs

As part of routine care, the nursing assistants administered a symptom questionnaire to the residents enrolled in the study and recorded residents’ vital signs daily, including blood pressure, pulse rate per minute, oxygen saturation and temperature.

#### Inclusion criteria

Residents of U.S. LTCFs where COVID-19 transmission was actively occurring. The LTCF medical director must agree to enroll the LTCF, and each participant must have the capacity to agree and sign consent.

#### Exclusion criteria

1) Current atrial fibrillation. NB: Paroxysmal atrial fibrillation is permitted if the participant is in atrial fibrillation less than 50% of the day on most days.
2) Pacemaker in place.
3) Known active infection other than COVID-19.
4) Dementia if the patient does not have a power of attorney in place.

#### Sample size and recruitment

For the current proposal, we selected two LTCF sites in the United States in real-time based on a prevalence of COVID-19 in the facility of 5-50%, which yielded a projected incidence of new cases in the LTCF in the ensuing 4 weeks of > 30%. Local and state departments of health and the VA regularly update the numbers of patients infected at each LTCF, so this information is readily available. In this report, we share the findings of the first 21 patients due to the urgency of the findings.

#### Follow-up duration

Total data collection for each participant is 60 days from the time of the first biometric recording.

#### Endpoints

Feasibility of biometric monitoring: Within 12 weeks we anticipated to prove that we can collect high-quality biometric data in the LTCF environment. Feasibility was assessed by reporting the proportion of quality signals, especially those capable of triggering an actionable alert, obtained out of all monitoring time for each device and compared to the frequency of such alerts triggered by the standard of care routine manual vital sign checks.

Algorithm development: We sought to identify biometric-derived features suitable for a statistical algorithm for the early detection of respiratory viral infections prior to deterioration and death.

#### Statistical analysis

A two-sided t-test for unequal variances was performed to compare the two groups of patients discharged to home or into palliative care. The number of diseased patients (n=4) precluded statistical comparisons, so we present these findings as clinical cases. F test was used to assess differences in variance. Differences were considered statistically significant for p<0.05.

## Results

From 12/2020 - 03/2021, we approached 26 PCR-confirmed SarsCoV2-positive patients at two SNFs: 5 declined, 21 were enrolled into monitoring by both sensors (female=13, male=8; age 77.2±9.1). One patient with arthritis discontinued the wearable due to wrist discomfort. We recorded outcomes as discharged to home (8, 38%), long-term care (9, 43%) or death (4, 19%). The O2Sat threshold of 91% was considered as hypoxia threshold.

Biostrap captured hypoxic events below 91% nine times as often as the routine intermittent pulse oximetry (**Figure 2**).

**Figure 2.**
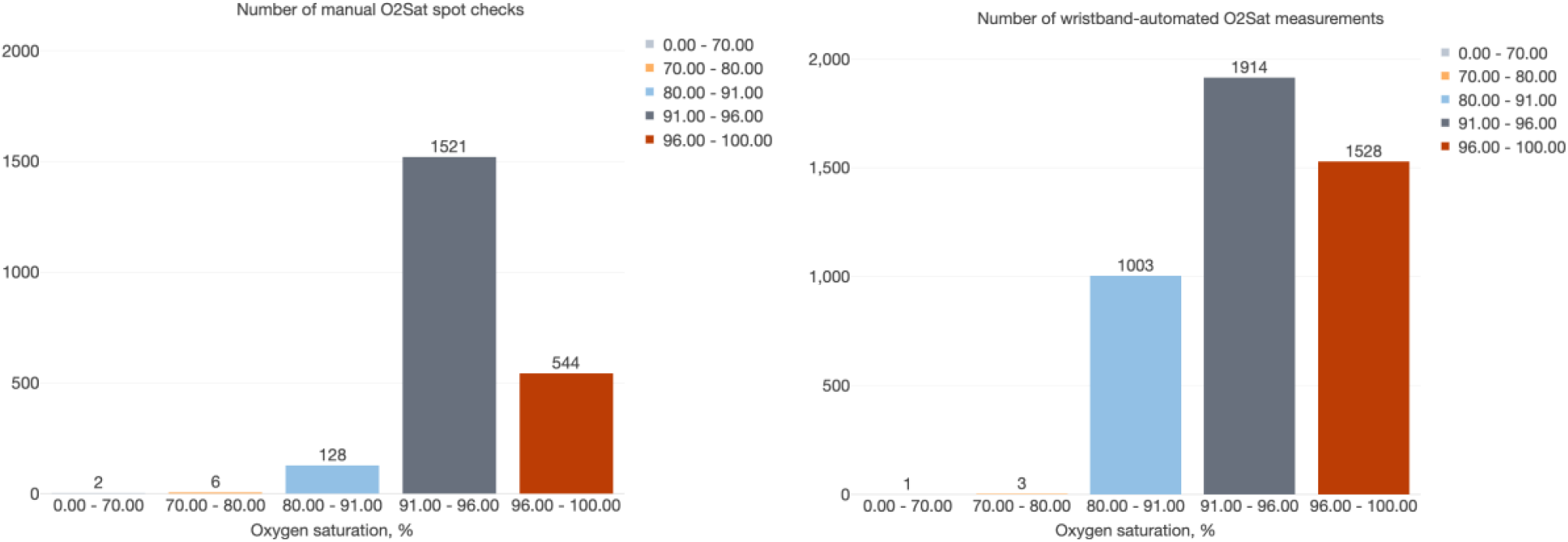
The ability to capture hypoxic events is markedly improved with wearable continuous O2Sat monitoring technology compared to the routine intermittent pulse oximetry monitoring. For senior patients diagnosed with COVID-19 and treated in the long-term post-acute care facilities, the threshold of 91% O2Sat is used as the alerting threshold triggering intervention.

Among the patients discharged to long-term care, a total of 76 days were monitored, HR did not differ compared to the patients discharged home (68±8 vs 70±7 bpm). However, we observed a statistically significant reduction of RR at 16±4/min as well as the variances in RR of long-term care patients vs. patients discharged home (**Figure 3**).

**Figure 3.**
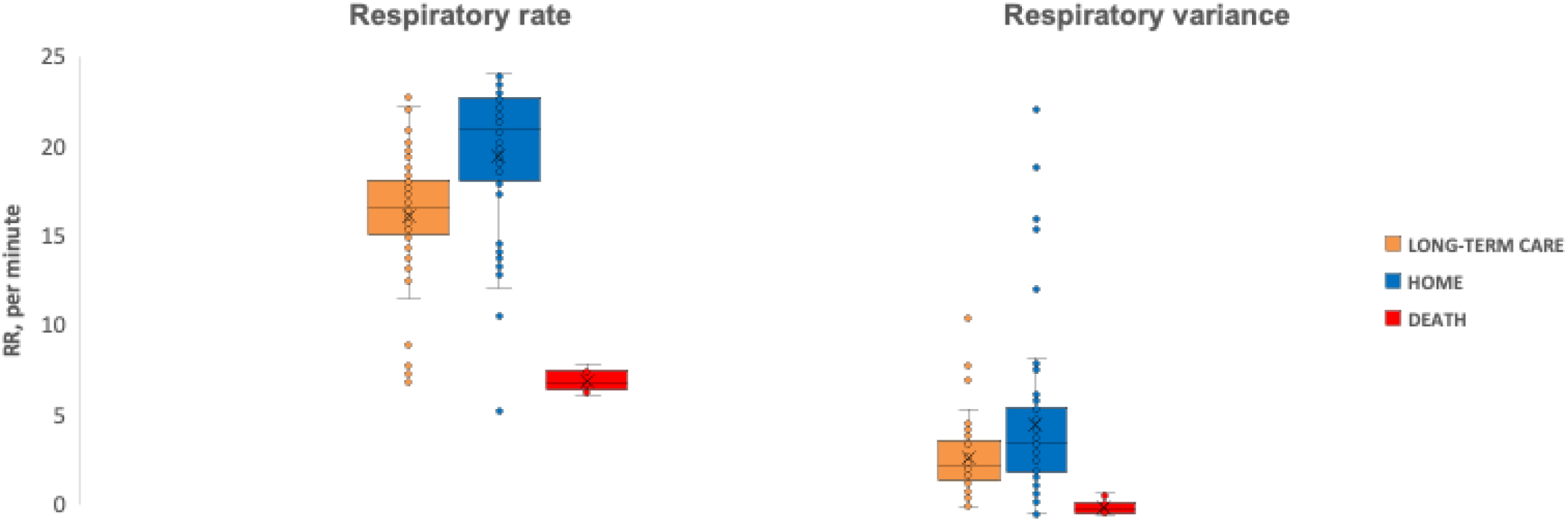
Under-mattress BCG sensor captured alterations in respiratory rate (RR) and RR variance over the period of stay.

In one patient deceased, two weeks prior to death, we observed a wide range of O2Sat values (65-95%) captured by Biostrap device and not noticeable with the routine vital sign spot checks (**Figure 4**). In this patient, the Emfit sensor yielded a markedly reduced RR (7/min) in contrast to 18/min from two routine spot checks performed in the same period of observation (**Figure 3**). This difference in RR also applied when compared the deceased patient to the seven patients discharged home over a total of 86 days of monitoring (RR 19±4) (**Figure 3**).

**Figure 4.**
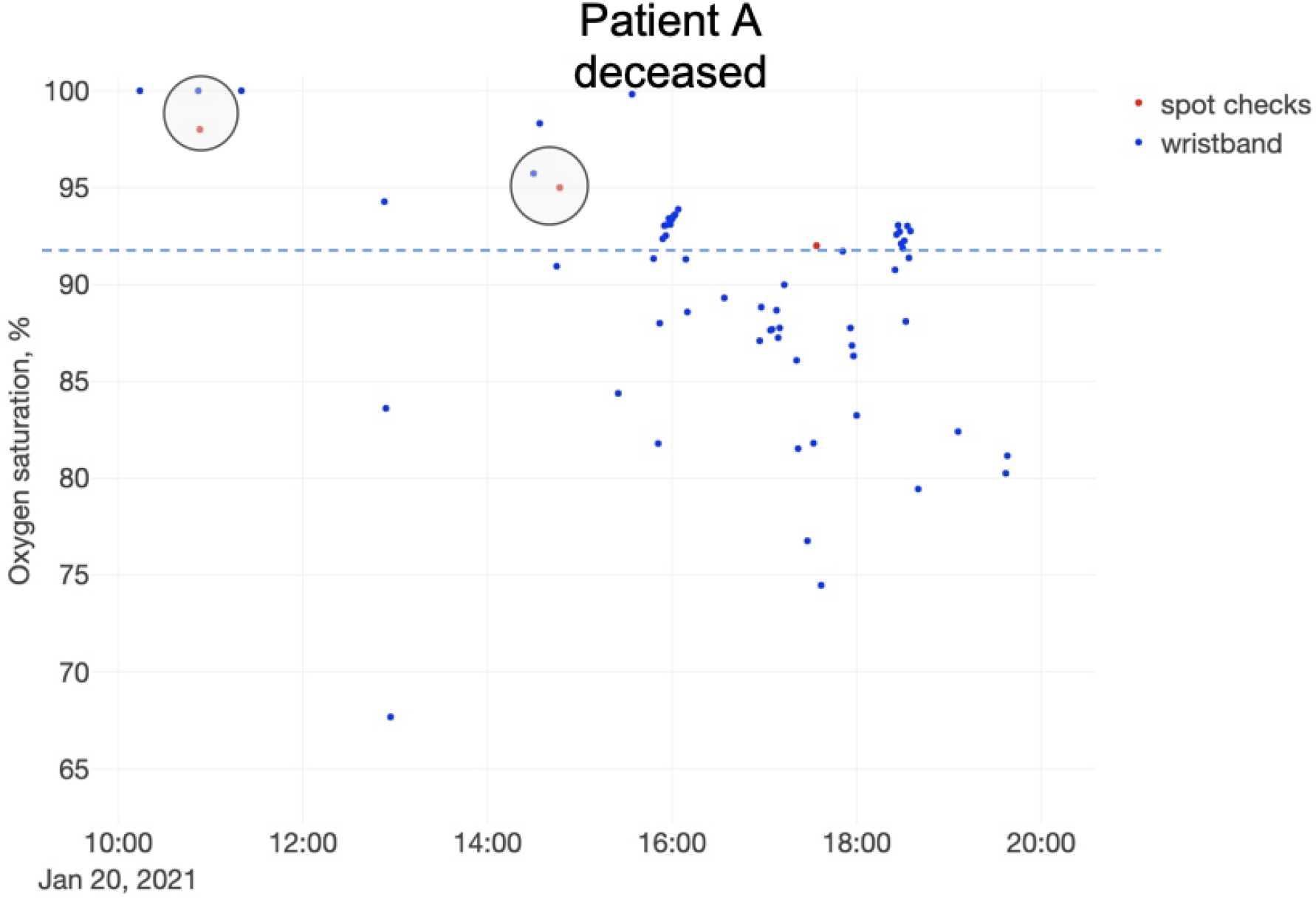
O2Sat measurements by the wristband wearable (blue) and routine vital sign spot checks (red). Note the close correspondence between both types of measurements where they happen closely together, and the intermittent severe hypoxic episodes captured by the wristband.

While the biometrics presented above are physiologically similar to those obtained intermittently manually (HR and RR), the variance of RR is a not a conventional metric of RR; it is uniquely accessible when continuous monitoring is enabled, such as presented here.

Next, we present the findings of another unconventional metric of physiological health in this population, the movement activity and variance of activity during sleep captured by the under-mattress sensor. While sicker patients spend more time in bed, the relative daily average and variance of movements while in bed are not affected by this fact. In the following, we report the statistical differences in these movement metrics between the patients discharged home and those who were discharged into long-term care. Because the sample size of patients who died did not permit statistical analysis, we compare the data we obtained as individual cases against the background of the values distribution found in both other groups.

Over the course of the entire monitoring period, among the seven patients discharged to long-term care, we observed a reduction of the variances in movement (but not in activity per se) compared to those who returned home (**Figure 5**). In the patient who died, intuitively, we observed stark reductions in mean activity and variances of activity in contrast to the patients discharged home.

**Figure 5.**
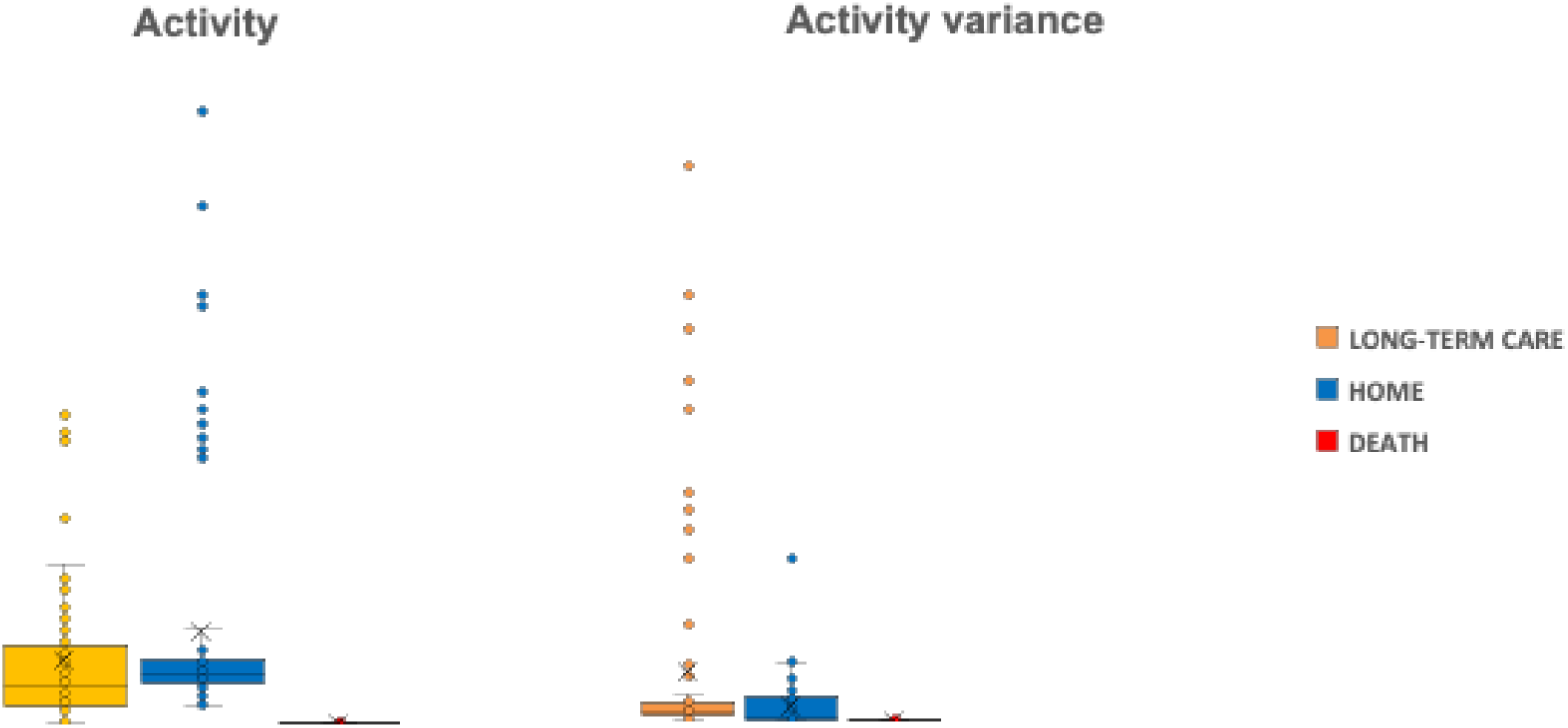
The under-the-mattress sensor captures differences in activity and its variance in senior post-acute care patients with COVID-19 discharged to long-term care compared to those discharged home or deceased. Note the reduction in activity variance (but not activity per se) in patients discharged to long-term care compared to those returning home.

## Discussion

Wrist-worn wearables and contact-free under-mattress sensors can be integrated successfully into the SNF workflows and are well tolerated by the patients. Interestingly, patients who were discharged to long-term care had a lower RR, possibly due to known neurotropic effects of SaRS CoV-2.[7] These effects are also likely the cause for the observed severe hypoxic periods in the deceased patient when using continuous O2Sat monitoring. Specific early changes of oxygen saturation fluctuations and RR heralded deterioration from COVID-19 two weeks in advance of standard manual O2Sat measurement and evaded timely detection without the device.

Our findings regarding the activity patterns indicate monitoring sleep quality can yield additional predictive insights into the patients’ outcomes in SNF setting. Future studies should explore the ability of wearables and contact-free sensors to provide predictive analytics from capturing sleep quality metrics such as tossing and turning as well as duration of sleep stages (REM, NREM).

### Reduction of viral transmission through detection of infection in asymptomatic stage

Detecting initial infection in time to effectively limit person-to-person viral transmission is impossible due to the long asymptomatic period (2-14 days) during which the individual, unaware that he or she carries the infection, continues to have contact with others. Health authorities and researchers have estimated that 30% of new SaRS CoV-2 infections are acquired through contact with asymptomatic people.[8] Solving the problem of detecting asymptomatic carriers who can transmit infection is key to protecting vulnerable residents of nursing homes and assisted living facilities, to protecting frontline workers who care for them, and to facilitating return to work (including the return of nurses and medical assistants).

Future studies should validate our algorithms in the pre-COVID stage setting; that is, in the setting where medical directors and nursing staff caring for the patients will be alerted to the likely acquisition of a respiratory virus (prior to the onset of symptoms), and thus be able to promptly arrange confirmatory virus testing and strict quarantine of the affected resident (or coverage for the affected employee, who would stay home) such that transmission of SaRS CoV-2 and other respiratory viruses among residents and staff will be curtailed and possibly eliminated.

Based on the findings in the general population [5,6] and the encouraging results in the current study, we expect that the presented continuous monitoring approaches will be able to detect biomarkers of infection prior to clinical symptom presentation. In future work, we will show that isolating asymptomatic and pre-symptomatic virus carriers detected through biometric monitoring will reduce the incidence of new clinical cases by 25% to 80% compared to that projected or to homes where no such monitoring is deployed. If we successfully detect newly acquired infection early enough, continuous biometric monitoring and self-quarantine of suspected carriers may replace mass population quarantine, facilitating economic recovery.

### Reduction of ICU utilization and death rate

We expect that continuous biometric monitoring will allow clinicians to direct their attention to the patients who need them the most, to avoid discharging patients with mild infection who are likely to succumb to COVID-19 later, and administer treatment in a timely fashion to prevent deterioration. These impacts will be amplified in the underserved and underrepresented populations such as SNF residents.

### Continuous vital signs and physiological systems monitoring: practical deployment scenarios

The devices tested in this study provide comfortable, hassle-free health monitoring. Nursing homes and assisted living facilities can particularly benefit from such technology because of:

- high incidence of COVID-19 infection and deterioration, and
- existing infrastructure to ensure the wristbands are worn continuously and, together with under-mattress sensors, sending the data automatically to the server for continuous analytics.

During outbreaks or as an intermittent early warning prophylaxis, this approach can be combined with daily nasopharyngeal samples to classify patients as “infected” or “not” -- with COVID-19 or any of 15 other viruses -- taken at baseline, then biweekly and when clinically indicated, and cases of COVID-19 (or other respiratory viruses) and of deterioration are reported.

The applications and the benefits from using wearables and under-mattress health monitoring technologies are complementary and it is important to consider both for SNF use to ensure that a range of possible use case scenarios is covered, depending on individual preferences and requirements of patients and SNFs.

Both technologies provide continuous, accurate physiological time-series monitoring of heart rate, heart rate variability, respiration, and (in the case of the wrist-worn wearables) oxygen saturation using a comfortable, hassle-free biosensor. We demonstrated that these biosensors are easy to use in the SNF environment. There are some technical differences that may drive deployment choices. The wrist-worn device we tested synchronizes with a data hub (such as a smartphone), 1:1, remotely via Bluetooth and requires intermittent, daily or once a week, charging. The contact-free under-mattress sensor uses BCG technology, provides continuous automated data synchronization via WiFi. The charging requirement for the wearable is easily solved in the SNF setting, e.g., meal intake, while the under-mattress sensor is always plugged into power and requires no attention at all. All these aspects represent particularly important advantages in this era of pandemic and facilities lockdown when the contact with the residents is kept to an absolute minimum.

## Conclusions

For secondary prevention, changes in HR variability (HRV) have repeatedly shown promise in identifying non-invasively effects of infection or general morbidity. For example, HRV metrics distinguished children affected by previous, even intrauterine infections such as Zika virus, hospital readmission, inflammation (in specific organs and systemically), fetal hypotension or acidemia, deterioration in adult intensive care unit (ICU) patients before it becomes irreversible.[9–16] These algorithms will inform the design of algorithms aimed at detecting the onset of COVID-19 or other life-threatening infections and early, subclinical stages of physiologic decompensation.

For primary prevention, if widely disseminated among vulnerable populations and the community-at-large, wearable biometric technology and sensors built into the environment, e.g., under the mattress, will help avoid the ravages of seasonal flu and other contagious illnesses, and we will be better prepared for future waves of COVID-19 or other pandemics. Even as vaccines are rolled out, due to immune senescence and immunocompromise, or evolutionary mutations in the virus, elderly people and those with chronic medical conditions may not be as well protected by it. Continuous biomonitoring provides another layer of protection for them.

In summary, wearable devices and under-mattress sensors in SNFs hold significant potential for early disease detection and prevention.

## Data Availability

The data produced in the present study are available upon reasonable request to the authors and upon the IRB approval.

## Acknowledgments

The authors would like to thank the staff of Avalon Healthcare skilled nursing facilities for their tireless efforts in enabling the study under the most difficult circumstances of the COVID-19 pandemic. The key findings have been presented at the AMDA 2022, March 10-13, Baltimore, MD, USA.

